# Informed consent and trial prioritization for human subject research during the COVID-19 pandemic. Stakeholder experiences and viewpoints

**DOI:** 10.1101/2022.10.31.22281754

**Authors:** Stefanie Weigold, Susanne Gabriele Schorr, Alice Faust, Lena Woydack, Daniel Strech

## Abstract

**Background:** Very little is known about the practice-oriented challenges and mitigation strategies for effective and efficient translation of informed consent and study prioritization in times of a pandemic. This stakeholder interview study aimed to identify the full spectrum of challenges and mitigation strategies for informed consent and study prioritization in a pandemic setting.

**Methods:** We performed semi-structured interviews with German stakeholders involved in human subject research during the COVID-19 pandemic. We continued sampling and thematic text analysis of interview transcripts until thematic saturation of challenges and mitigation strategies was reached.

**Results:** We conducted 21 interviews with investigators, oversight bodies, funders and research support units. For the first topic informed consent we identified three main categories: consent challenges, impact of consent challenges on clinical research, and potential response strategies for consent challenges. For the second topic prioritization of trials, we identified two main categories: need for prioritization of clinical studies and potential response strategies for prioritization of clinical studies. All main categories are further specified with subcategories. A supplementary table provides original quotes from the interviews for all subcategories.

**Discussion:** Mitigation strategies for challenges with informed consent and study prioritization partly share common ground. High quality procedures for study prioritization, for example, seem to be a core mitigation strategy in dealing with informed consent challenges. Especially in a research environment with particularly high uncertainty regarding potential treatment effects and further limitations for valid informed consent should the selection of clinical trials be very well justified from a scientific, medical, and ethics viewpoint.

## Introduction

One of the most challenging part of a pandemic such as COVID-19 is the lack of evidence-based knowledge about the prevention, diagnosis and treatment of this new disease. In a very short time, therefore, a worldwide need for research is emerging. The COVID-19 pandemic resulted in thousands of clinical trials on the same disease being planned, reviewed, funded, conducted and published worldwide within a few months [1]. As of June 2020, more than 2300 trials were registered in the clinicaltrials.gov registry, and more than half of these were interventional trials. This extremely high number of clinical trials, combined with the intense time pressure, very uncertain evidence, and pronounced infection control measures, equally led to an amplification of many research ethics challenges and, in some cases, even very pandemic- or COVID-19-specific challenges [2]. From a legal and ethical perspective, it is undisputed that even in pandemic or other exceptional situations, research ethical standards must be adhered to in order to protect study participants [3]. Based on the our own survey results [2], a scoping literature review and informal discussion with key informants for human subject research on COVID-19 in Germany led to two topics that are of particular practical and ethical relevance: challenges with informed consent and challenges with prioritizing clinical trials.

The informed consent process shall allow eligible patients to make their own judgments about benefits, risks/burdens, and other relevant aspects that come with study participation. Based on these judgements the patients make informed decisions about whether to participate or not. Valid informed consent exists only if the patients concerned were a) competent to make decisions in the situation in question, b) received the relevant information, c) understood it, and d) were free from coercion to decide for or against consent. Critical care settings in general bear several challenges for ensuring a valid consent process due to difficult patient- and context-related circumstances that decrease decision making competence [3, 4]. A pandemic situation with social distancing measures in place and high uncertainty regarding potentially effective treatments can further aggravate these challenges.

At the same time the efficient and effective recruitment of eligible patients for clinical studies to investigate pandemic-specific prevention or therapy approaches is of utmost importance. Prioritization of clinical studies that is the ranking of more or less important trials might become relevant, for example, due to a shortage of eligible patients. If all planned trials recruit at the same time but too little patients are eligible, clinical trials might face recruitment failures or trialists compete in recruitment activities. Recent studies showed that many clinical trials, for example in Germany, failed due to insufficient recruitment [5]. Prioritization might also be important if too many studies are conducted in an inefficient or even harmful way. For example, studies showed that many trials investigated the same therapeutic approach at the same time [6].

While some conceptual papers discussed COVID-19 specific challenges for informed consent [7] and study prioritization [8], very little is known about the practice-oriented challenges and mitigation strategies for effective and efficient translation of informed consent and study prioritization in times of a pandemic. The objective of this stakeholder interview study, therefore, was to identify the spectrum of challenges and mitigation strategies for *informed consent* and *study prioritization in a* pandemic setting. The results of the interview study shall inform the development of practice-oriented guidance for informed consent and trial prioritization as a component of pandemic preparedness.

## Methods

Ethical Approval: All steps of analysis in this study involving human participants were in accordance with the ethical standards of the research ethics committee of Charité Berlin (Approval No: EA4/006/21) and with the 1964 Helsinki Declaration and its later amendments.

Participants: We aimed to interview key stakeholders involved in human subject research during the COVID-19 pandemic until thematic saturation is reached. Based on typical sample sizes in qualitative research and own experiences we estimated that 15-20 interviews are needed to reach thematic saturation on the level of major and first-level subgroup categories. Key stakeholders include clinical investigators, clinicians from clinical trial centers who deal with COVID-19 patients on a daily basis (intensive care and normal ward), clinic directors, patient representatives as well as members of research ethics committees, of regulatory bodies, of public sponsors, of guardianship courts and authorities, and. The recruitment of interviewees followed a purposive sampling approach.. We identified representatives of the above mentioned stakeholder groups via the network of the QUEST Center, the AKEK (German working group of research ethics committees), and the PRECOPE advisory board. We further identified potential interview participants via snowballing.

Procedure: All potential interview participants received an email including study information (supplementary table S1) and the informed consent document (supplementary table S2). All participants provided written informed consent. We conducted the interviews via videoconferencing between March 2021 and July 2021. Interviews lasted between 30 and 60 minutes. One team member led the interview (LW, SW or DS, all trained in qualitative research methods), while another team member observed and made notes on its process and content (LW, SW, DS or AF). All interviews were guided using a semi-structured topic guide (supplementary table S3). It focused on challenges and strategies for *informed consent* and *prioritization in the context of Covid-19 trials* and was pilot tested with researchers of our institution. After each interview, the team conducted a peer-debriefing. A transcription company transcribed the interviews under a confidentiality agreement. MAXQDA 2020 was used for the analysis of the interview material.

Data analysis: We used qualitative content analysis according to Mayring (2010) [9]. The analysis consisted of an inductive development of thematic categories. The transcripts were coded by SW and 10 transcripts were verified (second coding) by AF. All codes and discrepancies within the coding were discussed and further modified within the research team (SW, LW, AF, SGS, DS) until a consensus was reached.

## Results

### Demographics

We conducted 21 semi-structured in-depth interviews with 21 participants to reach thematic (qualitative) saturation. Altogether, we contacted 51 stakeholders, which gives a response rate of 42%. Our sample included physicians working in clinical research (principal investigators and non-principal investigators), heads of clinical departments that were responsible for the enrollment of patients with COVID-19 in clinical studies, representatives from clinical study centers, members of research ethics committees, and a public funder. For complete demographics see table 1.

**Table 1:** Demographic data of interview participants

| Category | Subcategory | Frequency (n) |
| --- | --- | --- |
| Sex | Female | 7 |
|  | Male | 14 |
| Age | 30-34 | 3 |
|  | 35-45 | 7 |
|  | 45-50 | 3 |
|  | 50+ | 8 |
| Function | Principle investigators (PI) of Covid-19 human subject research | 8 |
|  | Non-PI investigators | 1 |
|  | Physicians (at university hospitals) | 7 |
|  | Medical University-based clinical research coordinator | 1 |
|  | Members of research ethics committees | 3 |
|  | Member of public funder | 1 |

For the first topic *informed consent* we identified three main categories: consent challenges, impact of consent challenges on clinical research, and potential response strategies for consent challenges. For the second topic *prioritization of trials*, we identified two main categories: need for prioritization of clinical studies, potential response strategies for prioritization of clinical studies. All main categories were further specified with first and second level subcategories (see table 2 for an overview). In the following we give a narrative overview of the identified main and subcategories. The corresponding original quotes from the interviews can be found in supplementary table S4

**Table 2:** Qualitative spectrum of topics regarding the informed consent for or prioritization of clinical studies on Covid-19.

| **Informed Consent**<br>Main categories and first-level sub-categories | Second-level sub-categories | Code |
| --- | --- | --- |
| Limitations for consent |  |  |
| Time pressure | Rapid deterioration of the patient's condition | C1 |
|  | Reduction of contact time | C2 |
| Isolation/hygiene conditions | Difficulty in building the doctor-patient relationship | C3 |
|  | Need to reduce the number of patient contact | C4 |
|  | Restricted accessibility to legal guardians due to quarantine regulations | C5 |
|  | Contact restrictions for relatives | C6 |
| Overburdening the patient | Distress due to parallel/competitive study recruitments | C7 |
|  | Difficulties in understanding due to complexity | C8 |
|  | Restrictions of the patients due to severe symptomatology | C9 |
|  | Increased risk of therapeutic misunderstanding | C10 |
| Negative impact on clinical research |  |  |
| Heterogenous ethics review |  | C11 |
| Delay |  | C12 |
| Recruitment failure |  | C13 |
| Bias | Excluding critically ill patients introduces important bias | C14 |
| Potential response strategies |  |  |
| Opt-in consent as ethical standard | Relevance for interventional studies | C15 |
|  | Conflicts of interest | C16 |
| Secondary-use of patient data | Broad consent model | C17 |
|  | Opt-out model | C18 |
|  | No consent model | C19 |
| Patients incapable to give consent | Need of consistent IRB requirements | C20 |
|  | Deferred and legal proxy consent | C21 |
|  | Alternative proxy decision makers | C22 |

| **Study Prioritization**<br>Main categories and first-level sub-categories | Second-level sub-categories | Code |
| --- | --- | --- |
| Need of prioritization |  |  |
| Patient protection | Prioritization could improve risk-benefit assessment for clinical trials | P1 |
|  | Prioritization could prevent overburdening of patients | P2 |
| Validity and efficiency | Prioritization could reduce the high number of parallel/redundant studies | P3 |
|  | Prioritization could improve quality of studies | P4 |
|  | Prioritization could prevent trials suffering of insufficient resources and expertise | P5 |
|  | Prioritization could reduce inefficiencies due to competitive recruitment | P6 |
| Relevance | Prioritization could support the relevance of clinical studies | P7 |
|  | Prioritization could identify patient-oriented knowledge gaps | P8 |
|  | Prioritization should include the allocation of biomaterials | P9 |
| Lack of explicit prioritization |  | P10 |
| Potential response strategies for prioritization |  |  |
| Central coordination and collaboration | National board | P11 |
|  | National cooperation | P12 |
|  | Central registry | P13 |
|  | Consultation by Ad-hoc expert groups | P14 |
|  | Efficient study planning | P15 |
|  | Effective recruitment of special patient groups | P16 |
|  | Management of conflict of interest | P17 |
| Study design | Multicentre study design | P18 |
|  | Adaptive study designs | P19 |
| Alignment with funding | Funding for priority studies | P20 |
|  | Calls for priority topics | P21 |
|  | Efficient funding procedures | P22 |
|  | Fair funding procedures | P23 |
|  | Sustainable funding | P24 |
| Pandemic policy on use/ownership of patient data | Agreement for secondary data use | P25 |
|  | Regulation on data use and access | P26 |
| Professionalisation of clinical research | Competence in clinical research | P27 |
|  | Fair play among scientists | P28 |

### Informed Consent

#### Consent challenges

##### Time pressure

Various interviewees emphasized that they needed to obtain the informed consent under increased time pressure. Often this was due to the ‘rapid deterioration of the health condition of patients’ with COVID-19 (C1). Furthermore, interviewees indicated the need to ‘reduce contact time’ to minimize the risk of infection (C2).

##### Isolation and hygiene conditions

Various interviewees indicated ‘difficulties in building the doctor-patient-relationship’ due to the intended contact restrictions and the protective clothing (C3). Within the strict isolation conditions, the ‘number of patient contacts’ and the duration of these were reduced in order to minimize the spread of the virus (C4). In addition, it was highlighted that the ‘reduced accessibility to legal guardians’ (C5) and the ‘contact restrictions for relatives’ (C6) had negative impact on informed consent procedures.

##### Overburdening the patient

Interviewees reported several aspects of the pandemic situation that carried a risk to overburden the patient and thus weaken the quality of the informed consent procedure. Some patients were confronted with ‘parallel/competitive study recruitment’ which could cause distress at the patient side (C7). Patients often had difficulties in ‘understanding due to complexity’ of informed consent documents. This was further complicated by the pandemic context (C8). The informed consent procedure could especially overburden those patients that already suffered ‘severe symptomatology’ and rapid disease progression (C9). Furthermore, some interviewees mentioned an ‘increased risk of therapeutic misunderstanding’ when the disease is rather novel (C10).

#### Impact of consent challenges on clinical research

The challenges around appropriate consent procedures for COVID-19 trials had an impact on conducting clinical research:

##### Heterogenous ethics review

Some interviewees mentioned ‘heterogenous ethics review’ (C11). They referred to an IRB decision that forced to exclude those trial participants that initially gave consent but loose the decision making competence over the course of the trial due to disease progression. Other IRBs allowed the retention of these trial participants.

##### Delay and recruitment failure

Interviewees pointed to the potential ‘delay’ (C12) of conducting trials or even ‘stops/recruitment failures’ (C13) that result, for example, from difficulties to contact legal proxies during a pandemic.

##### Bias

Interviewees further pointed to the risk that even in conducted studies the exclusion of critically ill patients due to consent limitations would ‘introduce important bias’ (C14).

#### Potential response strategies for consent challenges

##### Opt-in consent as ethical standard

Despite the challenges experienced, several interviewees emphasized the *informed consent* and the self-determination of patients as the ethical standard with particular ‘relevance for interventional studies’ (C15). In addition to patient-centered considerations, some interviewees addressed the protective function of the informed consent for potential ‘conflicts of interest’ of clinician investigators (C16).

##### Secondary-use of patient data

Regarding non-interventional studies, various interviewees suggested the implementation of a ‘broad consent model’ for the use of routine data and biomaterials (C17). The opportunity to learn immediately from routine care with legitimate access to routine data was emphasized. By pointing to the potentially high social value of secondary use of patient data during a pandemic interviewees suggested an ‘opt-out model’ (C18) and a discussion on when even a ‘no consent model’ for the secondary use of patient data might be appropriate (P19).

##### Patients incapable to give consent

When discussing how to better prepare for potential future pandemics nearly all interviewees expressed the need to implement consent strategies for those patients who are incapable to give consent. Interviewees highlighted the ‘need of consistent IRB requirements’ especially for study participants who lose their consciousness after prior consent (C20). Several interviewees described the ‘deferred and legal proxy consent’ (C21) as an appropriate option. However they referred how difficult and challenging it is for relatives to make such a proxy decision on trial participation especially in a pandemic situation. Some interviewees mentioned the option of ‘alternative proxy decision maker’ (C22) such as physicians or ethics committees.

#### Prioritization aspects

##### Need of prioritization

###### Patient protection

Interviewees described that if prioritization of clinical trials could help reduce unnecessary risks that might come with participation in less well justified trials this would support patient protection and thus ‘improve risk-benefit assessment’ of planned clinical trials (P1). Prioritization could also ‘prevent the overburdening of patients’ that are asked from different physicians at the same time to participate in their clinical trials (P2).

###### Validity and efficiency

Interviewees mentioned the need to ‘reduce the number of parallel/redundant studies’ that were conducted during the COVID-19 pandemic (P3). Several interviewees further highlighted that the designs of COVID-19 trials often lacked measures to increase the robustness and that the reporting quality of study protocols was low. A more explicit prioritization process might therefore ‘improve the quality of clinical trials’ (P4) and could ‘avoid that trials are conducted with insufficient resources and expertise’ (P5). As above mentioned, in some occasions one patient was asked to participate in several studies illustrates the benefit of prioritization measures to ‘reduce inefficiencies’ due to competitive recruitment (P6).

###### Relevance

Numerous interviewees pointed out that several research questions addressed in COVID-19 studies lacked relevance. Prioritization could therefore ‘support the relevance’ of clinical studies (P7). Some interviewees commented on important criteria for prioritization and most of them mentioned clinical relevance as a top criterion. Some interviewees criticized that most studies investigated different drug treatments and only few procedures for COVID-19 such as lung ventilation. Therefore, more explicit prioritization could not only select among already planned studies but also ‘identify patient-oriented knowledge gaps’ (P8). Interviewees mentioned that prioritization also becomes relevant in ‘allocating the scarce resource of biomaterials’ such as lung tissue from patients with COVID-19 (P9).

###### Lack of explicit prioritization

Our study did not aim to gather quantitative information about how often our interview participants experienced explicit prioritization on which trials to conduct and which not. We want to highlight, however, that from all 21 interviewees engaged in conducting or coordinating clinical research all except one reported that they did not experience any type of explicit prioritization of clinical studies. Several interviewees stressing this ‘lack of prioritization’ at the same time highlighted the different needs for prioritization as presented above (P10).

#### Potential response strategies for prioritization

##### Central coordination and collaboration

Interviewees repeatedly emphasized that central coordination via, for example, a ‘national board’ (P11) and ‘national cooperation’ (P12) could be helpful to prioritize clinical studies. The RECOVERY trial [10] and other WHO studies were mentioned as having created relevant evidence via respective coordination and collaboration efforts. In order to profit from the specific expertise and study ideas at individual university hospitals interviewees proposed a ‘central registry’ (P13). Another interviewee suggested the development of a procedure for ‘ad-hoc expert groups’ (P14). This ad hoc expert groups should be set up for specific urgent topics with members from different institutes that jointly discuss and design a specific study. Interviewees highlighted that central coordination could also improve ‘efficiency in study planning’ (P15) on a national level that might allow Germany to better participate in international activities such as the WHO studies. Furthermore, central coordination was also mentioned as a way to support ‘effective recruitment of special patient-groups’ (P16). Additionally, interviewees highlighted that the ‘management of conflict of interests’ could be supported by more central coordination and collaboration (P17).

##### Study design

Interviewees mentioned that study designs like ‘multicenter study’ (P18) or ‘adaptive study designs’ (P19) could be helpful to improve recruitment and efficiency.

##### Alignment with funding

While discussing the issue of prioritization different interviewees mentioned a lack of public calls for therapeutic studies. Prioritization should align with opportunities for ‘funding priority studies’ (P20) and ‘calls for priority topics’ (P21). Effective prioritization is further dependent on ‘efficient funding procedures’ (P22) and ‘fair funding procedures’ (P23). The fairness of funding was also contextualized as a consequence of more explicit prioritization if the relevance of certain research questions receives more priority than for example the size of a university. ‘Sustainable funding’ (P24) becomes important to make the prioritization procedures effective.

##### Pandemic policy on use/ownership of patient data

Several interviewees pointed out that a strong ‘agreement for secondary use of data and biomaterials’ is needed for an effective and fair prioritization in the secondary use of patient data and biomaterials (P25). To support this agreement a ‘regulation on data use and access’ is one important aspect that should be clarified upfront in preparing for pandemics (P26).

##### Professionalization of clinical research

In elaborating strategies and success conditions for prioritization of clinical studies several interviewees pointed out that more ‘competence in clinical research’ (P27) is needed in Germany. During the pandemic, sometimes people with insufficient training in study design and other clinicalvresearch related competencies organized and conducted clinical research. Moreover, interviewees pointed out that the need for ‘fair play among scientists’ (P28) serves as a condition of success for collaborative and effective research that qualifies for prioritization.

## Discussion

This qualitative interview study investigated stakeholders’ experiences and viewpoints regarding informed consent and trial prioritization for human subject research during the COVID-19 pandemic.

### Informed consent

For the topic of informed consent the interviewees mentioned significant practical limitations that can negatively impact on the conditions for a valid consent. The competence to make autonomous decisions and the understanding of the relevant information can be impacted by practical limitations such as “time pressure” or “isolation conditions”. Further challenges such as competitive recruitment might “overburden patients” and thus impact on competence and understanding. The often high uncertainty about expected benefits and harms of investigated treatments (over 99% of investigated COVID-19 treatments failed [11]) also impact on the “relevant information” condition for a valid consent. The more the validity conditions for informed consent are challenged the less does the consent procedure serves its role of patient protection. Our qualitative study cannot provide an answer to the question of how often obtained consent in COVID-19 studies was invalid. The plausibility of having strongly limited conditions for valid consent during a pandemic urgency situation, however, signals the need to define and implement measures that either facilitate better validity conditions or compensate for the decreased protective function of informed consent. We come back to these compensating measures below.

The challenges around informed consent cannot only affect the protection of trial participants but can also limit the effective and efficient conduct of clinical research and thus affect the society at large. Relevant factors potentially limiting clinical research are recruitment challenges (via delay or failure of studies), biased sampling of less severely affected patients (that have higher chances to give consent in a timely fashion). Here again, our qualitative and thus explorative study can only raise awareness about these challenges but we cannot specify how often specific trials were strongly delayed or failed due to consent challenges. Data on recruitment failures in Germany point to relatively low recruitment rates and a lot of early stopping of trials [5]. The challenges of informed consent could be one contributing determinant beside others. To prepare for future pandemics with maybe even more challenging conditions for valid informed consent procedures further ethical, legal and social discussions including all relevant stakeholder groups should address the question of what modification of consent procedures are acceptable in pandemic emergency situations that strongly require clinical research especially with severely affected patients that are not able to give consent.

Our interview results suggest that secondary use of patient data under a broad consent model or even an opt-out model might resolve some of the above mentioned challenges. An exemplary discussion for Low and Middle Income Countries (LMIC) can be found at Singh et al. (2021), who evaluate how conducting biobank research under a waiver of informed consent during public health emergencies is ethically permissible [12]. However, while secondary use of health data can create additional information, it cannot replace clinical trials.

In contrast to Germany, in other countries alternative *consent strategies* were considered legitimate under the special circumstances of the pandemic. Repeatedly, interviewees referred to the knowledge gained rapidly by the Recovery Study conducted in the UK [10]. To be emphasized is the *IC* agreement, already anchored in the study protocol, which made deferred consent solutions possible by a clinician in consultation with the research ethics committees. Patients who lack capacity to consent due to severe disease (e.g. need ventilation), and for whom a legally designated representative is not available, randomization and consequent treatment will proceed with consent provided by a clinician independent of the clinician seeking to enroll the patient who will act as the legally designated representative. Consent will then be obtained from the patient in case of recovery or the patient’s personal legally designated representative at the earliest opportunity [10].

### Prioritization aspects

When reflecting the urgent need to generate knowledge and at the same time the need to select the most promising studies and avoid wasteful redundancies and inefficiencies in clinical trials, interviewees emphasized the potential value of well-informed prioritization decisions. The common task would be to decide which (often collaborative) trials to launch and to assure that they are robust and create patient-oriented output. A successful, frequently cited example is the already mentioned centrally planned multicenter RECOVERY study [10]. Organized and planned by Oxford University, it was initiated with the National Health System. It recruited 10,000 patients within two months and ultimately produced substantial evidence for the treatment of COVID-19 (ibid.) [13].

With regard to the German context, interviewees saw the German Network University Medicine [14] which was founded during the pandemic to foster cooperation between UMCs, as a fruitful starting point. The network aims to bring together and evaluate action plans, diagnostic and treatment strategies from as many German university hospitals as possible. This bundling of competencies and resources is intended to create structures and processes in the hospitals that ensure the best possible care for COVID-19 patients. In addition, interviewees discussed that non-university institutions and networks established through the Robert Koch Institute, for example, should also be considered as collaboration partners. Within all these institutions, databases and registers have been created in preparation for evidence synthesis in future pandemics, which are helpful for potential decision-making (e.g. CEOsys [15], NAPKON [16], CODEX [17] [18], STAKOB [19]).

A differentiated approach for possible prioritization criteria was also developed by Meyer et al. [8] who have drafted guidance for research institutions on how ethical consolidation and prioritization of COVID-19 clinical trials might proceed in a three-stage assessment. It first considers whether a study meets the criteria of social value, scientific validity, feasibility, and collaboration. Second, if all threshold criteria are met, the institutional capacity to conduct a study should be examined and assessed. Third, study-specific prioritization criteria should be evaluated, concerning the safety and effectiveness of interventions, the robustness of designs as well as institutional resource utilization, expertise and experiences in clinical practice. Meyer at al. suggest further to check several diversity criteria, which query e.g., several patient life stage and risk factors, a wide range of disease stages, and ensure inclusions from different geographic and sociodemographic regions. For implementing these criteria, “COVID-19 research prioritization process should occur prior to IRB, grants office, and other institutional reviews to avoid wasting those important resources” [8]. In addition, communication of potential prioritization decisions as well emergency risk communication should be considered [20].

Because prioritization of specific studies can not only be helpful for scientists by improving quality but also protect patients by increasing the chance that primarily robust and clinically highly relevant studies are initiated. Prioritization and collaboration efforts to improve the scientific justification of trials conducted in the pandemic context would this function as an important compensation for the above mentioned challenges with the informed consent procedure.

### Limitations of the study

Our study has the following limitations. Firstly, the content focus on informed consent and study prioritization was set by us. As mentioned in the introduction, this was justified by literature-based preliminary work and expert consultation. Secondly, while we were able to include diverse perspectives from trialists, health care professionals, regulatory bodies we were not able to identify patient representatives that could comment more generally on experiences with informed consent and study prioritization in the pandemic context. We believe that a quantitative survey of trial participants and/or their legal representatives would be an important follow-up work on our explorative qualitative expert interviews.

## Conclusion

This interview study identified a broad spectrum of challenges and mitigation strategies for *informed consent* and *study prioritization in a* pandemic setting. Mitigation strategies partly share common ground. Procedures for study prioritization, for example, seem to be a core mitigation strategy in dealing with informed consent challenges. Especially in a research environment with particularly high uncertainty regarding potential treatment effects and further limitations for valid informed consent should the selection of clinical trials be very well justified from a scientific and practical relevance viewpoint. The results of the interview study shall inform the development of practice-oriented guidance for informed consent and trial prioritization as a component of pandemic preparedness.

## Data Availability

All data produced in the present work are contained in the manuscript. Full interview transcripts cannot be shared publicly because of participants privacy. Excerpts of the transcripts relevant to the study are provided within the paper (see supplementary table S4).

## Acknowledgement

We would like to thank the participating interviewees for their time and willingness to talk to us. We would like to thank further members from our research group for comments on the manuscript prior to submission.

## Availability of data and materials

Full transcripts cannot be shared publicly because of participants’ privacy. Excerpts of the transcripts relevant to the study are provided within the paper (see supplementary table S4).

## Author’s contributions

All authors participated in the conceptualization of the interview study and developed the interview guide. SW and LW conducted the interviews and were supervised by DS. SW, AF, DS and LW analyzed and interpreted the data. LW and SW wrote the first draft of the manuscript. DS supported to revise the manuscript. All authors read and approved the final manuscript.

## Ethics approval and consent to participate

All interviewees provided written consent to participate and the research ethics committee of Charité - Universitätsmedizin Berlin approved our study (Approval No: EA4/006/21).

## Supplementary Table S4

**Part “Informed Consent”:**

**Qualitative spectrum of topics with relevance for the informed consent for clinical studies during the Covid-19 pandemic**

| Main categories and first-level sub-categories | Second-level sub-categories | # | Quotes from anonymized interview transcripts |
| --- | --- | --- | --- |
| Limitations for consent |  |  |  |
| Time pressure | Rapid deterioration of the patient's condition | C1 | If you are the only person who can include a patient in a study today, then it has to happen today, because maybe tomorrow the patient will no longer be able to give consent or it will be too late. (Interview 14, item 6)<br>The most important points are how to deal with patients who are incapable of giving consent, who may still be capable of giving consent at the time of study inclusion or study screening, but then lose their ability to give consent during the course of the disease due to a deterioration in their condition, because, for example, they require ventilation or intensive care and so on. (Interview 8, item 2)<br>So, being severely respiratorily restricted and hardly able to be kept alive is of course a situation where you don't really want to inform anyone, but you need the decision now (Interview 5, item 16). |
|  | Reduction of contact time | C2 | [...] when you educate patients on site, you want to get as little material in and out as possible. And of course you also want to reduce the contact time inside and the frequency of going in and out, or keep it as low as possible. (Interview 6, item 25) |
| Isolation/hygiene conditions | Difficulty in building the doctor-patient relationship | C3 | The particular challenge in the case of a highly contagious disease, and this does not have to be in the context of a pandemic, is of course that it is less easy to give the patient the impression that you will now sit down with him in peace and have a personal conversation. If I come to a patient fully disguised and he can only see my eyes through a plexiglass pane and otherwise sees nothing of me, then it is of course difficult to establish a trusting basis for a partnership discussion. (Interview 1, pos. 5) |
|  | Need to reduce the number of patient contact | C4 | And of course one also wants to [...] reduce the frequency of going in and out, or keep it as low as possible. (Interview 6, item 25) |
|  | Restricted accessibility to legal guardians due to quarantine regulations | C5 | And what has been considered or what is of course also a problem is now also in the context of the pandemic, that of course even if legal guardians are available, that they are of course quarantined by the pandemic or even in quarantine. (Interview 12, item 24) |
|  | Contact restrictions for relatives | C6 | And then, of course, the problem of not having relatives within reach, which was easier before the pandemic due to hygienic conditions - those were actually the challenges. (Interview 17, item 2) |
| Overburdening the patient | Distress due to parallel/competitive study recruitments | C7 | For example, doctor A goes to the patient and asks: 'Do you want to participate in the XY drug study?' The patient asks for time to think about it. During the reflection period, doctor B comes and clarifies for the next trial. And the patient thinks it's the same study and then says yes. Things like that can happen. (Interview 6, item 70). |
|  | Difficulties in understanding due to complexity | C8 | [...] and it's actually so complex by now that I'm always worried whether the person who is not an expert can really do something with the information or whether it's not just overwhelming that they have to sign it. (Interview 10, item 2).<br>The documents are often 10, 12 pages long. That's too long and they're presented in such a complex way that the patient can't understand them anymore. (Interview 4, item 26)<br>Many were also skeptical (regarding study participation in a foreign disease such as COVID-19) [...] they often were in a very bad condition and then overwhelmed by all the information you have to transmit quickly. (Interview 7, item 2). |
|  | Restrictions of the patients due to severe symptomatology | C9 | The greatest challenge now with regard to studies was simply first of all to obtain consent, insofar as the patients were already breathing quite strenuously and had difficulties just thinking about it calmly. (Interview 7, item 2)<br>Even if you now say: "I only take in patients who initially come to a normal ward and are still so healthy, in quotation marks, that they can be easily educated," there are still a few<br>who have shortness of breath or fear, are confused or whatever. (Interview 1, item 15) |
|  | Increased risk of therapeutic misunderstanding | C10 | [...] this situation of "I'm now coming to the intensive care unit and I'm afraid of what's coming up and what's going to happen" [is] threatening and then someone else comes along and says, "I can help you, but you have to sign for me once." That is, of course, extraordinarily difficult. (Interview 1, item 4) |
| Negative impact on clinical research |  |  |  |
| Heterogenous ethics review |  | C11 | For example, in our drug study we were told that only patients who are able to give consent may be included. That means that as soon as they [those who have already given their initial consent] are no longer capable of giving consent, or as soon as they are intubated, they are kicked out. (Interview 14, item 31)<br>The question is: What do I do with the patient who goes to the intensive care unit? The hospital ethics committee has said, "In the observational study, you can just leave him in, even if he's sedated and ventilated in the meantime, and then he can't withdraw his consent." The *state agency* says, or a person in the *state agency* says, "The moment the patient goes into the ICU, the patient has to be excluded from the study." Although, of course, you could just as easily say, "Well, he consented, he wants to stay in the study, even though he can't comment on it now." But because he can no longer withdraw, you have to exclude him. (Interview 1, item 15) |
| Delay |  | C12 | We still have an insane number of clinical studies and now we have the situation again: At the moment, none of the centers is recruiting very well, because there are hardly any more patients. We must not forget that if the protocols are complicated, the inclusion and exclusion criteria will not work. (Interview 9, item 34) |
| Recruitment failure |  | C13 | We still have an insane number of clinical studies and now we have the situation again: At the moment, none of the centers is recruiting very well, because there are hardly any more patients. We must not forget that if the protocols are complicated, the inclusion and exclusion criteria will not work. (Interview 9, item 34) |
| Bias | Excluding critically ill patients introduces important bias | C14 | [...] if someone is not capable of giving consent at the beginning of the admission: For these patients, in my opinion, you absolutely have to do everything to be able to include them in the studies, because otherwise you get an enormous bias if you leave out the seriously ill. Then, of course, the significance is much, much lower. (Interview 16, item 77) |
|  |  |  | That's where a completely different argument comes in: The moment you eliminate the seriously ill, i.e. when you discard the material, you introduce a bias, and then it is no longer representative at all - you need the seriously ill. (Interview 16, item 92) |
| Potential response strategies |  |  |  |
| Opt-in consent as ethical standard | Relevance for interventional studies | C15 | I think everyone would fight tooth and nail against saying that informed consent should be abolished (Interview 10, item 30).<br>Then, I think, with drug studies, there always has to be some kind of consensus: I think that's important (Interview 14, item 24). |
|  | Conflicts of interest | C16 | Under no circumstances [modify the consent in times of crisis]! Physicians always have conflicts of interest. (Interview 3, *protocol*, item 3) |
| Secondary-use of patient data | Broad consent model | C17 | We put a lot of emphasis on the fact that in these two cohort studies, in staff and patients, these are the two cohorts, the broad consent both... so that there were two separate things created. A filling of the biobank with broad consent and then a registry. That allowed us to be able to collect virtually all of the routine data that was generated. We made sure that patients signed correct consents, which then allowed for re-contact afterwards and so on. (Interview 16, item 43) |
|  | Opt-out model | C18 | Exactly with such a, let's say, medical or such an inclusion procedure and then, so to speak, with such an opt-out solution, I would not find it so odd for register studies. (Interview 14, item 24) |
|  | No consent model | C19 | Intervention study versus just data summary. And that's where I just come first to the question of having and using data from patients for later and also bio-materials. And I have to say, for this part my impression at Covid-19 was that the interest for the general public is so high that it would be helpful to define what is generally possible for the general public at a high level and does not require consent. (Interview 11, item 4) |
| Patients incapable to give consent | Need of consistent IRB requirements | C20 | The question is: What do I do with the patient who goes to the intensive care unit? The hospital ethics committee has said, "In the observational study, you can just leave him in, even if he's sedated and ventilated in the meantime, and then he can't withdraw his consent." The *state agency* says, or a person in the *state agency* says, "The moment the patient goes into the ICU, the patient has to be excluded from the study." Although, of course, you could just as easily say, "Well, he consented, he wants to stay in the study, even though he can't comment on it now." But because he can no longer withdraw, you have to exclude him. (Interview 1, item 15) |
|  | Deferred and legal proxy consent | C21 | Well, I would not necessarily demand deferred consent, but a combination of deferred consent and a consultant would perhaps make it better, yes; that someone from a different field actually has to look at it. I think in observational studies this is relatively uncritical and in interventional studies, I think it's also appropriate. (Interview 1, item 23)<br>[...] a husband or a wife and they have an acute event and have to go to the intensive care unit and have to be ventilated and could be in a study. Then the life partner usually has to sign. But he is completely overwhelmed with the current situation, so I would delegate more to the physicians for such intensive care studies, where I simply have the know-how - this is such complex medicine. (Interview 4, pos. 30) |
|  | Alternative proxy decision makers | C22 | [...] a husband or a wife and they have an acute event and have to go to the intensive care unit and have to be ventilated and could be in a study. Then the life partner usually has to sign. But he is completely overwhelmed with the current situation, so I would delegate more to the physicians for such intensive care studies, where I simply have the know-how - this is such complex medicine. (Interview 4, pos. 30) |
|  |  |  | And we have this now again and again with the Covid patients in the sense of - now there is the topic group benefit - in this pandemic, in this danger for the entire population, the ethics committee has decided, if someone is no longer capable of giving consent and no caregiver is present and if nothing speaks against it, because some relative says: "No.", then the ethics committee takes the responsibility, so that biomaterial can be absorbed for the research immediately when a patient reaches the clinic. (Interview 16, item 90)<br>Exactly, that [the inclusion of seriously ill non-consenting patients in clinical trials] worsens the scientific validity - that is a gross mistake in my eyes. And for that you need an ethics committee, you see? For that you need an ethics committee that allows you to do that. (Interview 16, pos. 96) |

**Part “Prioritization”:**

**Qualitative spectrum of topics with relevance for the prioritization of clinical studies during the Covid-19 pandemic**

Part "Prioritization": Qualitative spectrum of topics with relevance for the prioritization of clinical studies during the Covid-19 pandemic
| Main categories and first-level sub-categories | Second-level sub-categories | # | Quotes from anonymized interview transcripts |
| --- | --- | --- | --- |
| Need of prioritization |  |  |  |
| Patient protection | Prioritization could improve risk-benefit assessment for clinical trials | P1 | [...] And then came the reconvalescent plasma study. It is still running. And the impression is: [...] We make not only no 'benefit', but we prolong the intensive care stay. We rather make problems. And finally, piece by piece, the studies came here that showed: "It doesn't help there. It doesn't help there. [...] when do you stop a study like that? (Interview 11, item 31) |
|  | Prioritization could prevent overburdening of patients | P2 | [...] the coordination and prioritization of the research projects at the interfaces of the various wards and disciplines, [...] all colleagues [...] want to do interventional studies, five pharmaceutical companies email per day, want to do something. And it has to be said that all of this has to be coordinated somehow and a sensible research portfolio has to be created that does not overburden the patient. You can't have people running to the same patient ten times a day and saying, "I'm going to tell you about this study and that one." That would be dubious in the end. And to realize that the patient's resilience is finite, but that certain things, such as biomaterial, blood, which can be taken, are also finite for certain things and to embed them in a meaningful portfolio, this is also a challenge. (Interview 8, item 4) |
| Validity and efficiency | Prioritization could reduce the number of parallel/redundant studies | P3 | It's [inquiries to participate in or recruit for clinical trials] completely unstructured. You are offered something, you don't have anything, and then you just take that, and sometimes there are actually overlaps. So that's problematic: We have several studies that are aimed at the same patient population and that's a problem. (Interview 5)<br>In principle, everyone expressed their interests in some way, and in the end there was a lack of clarity: "How do you do it when everyone announces their interests? In the end, will it be the case that certain groups, who perhaps have some form of - let me put it this way - influence, then perhaps implement their study interests, or how does that work? And in the case of such a pandemic, it would indeed have been helpful if certain therapies or studies or even diagnostic studies had been seen as a priority. (Interview 5)<br>We still have an insane number of clinical studies and now we have the situation again: At the moment, none of the centers is recruiting very well, because there are<br>hardly any more patients. We must not forget that if the protocols are complicated, the inclusion and exclusion criteria will not work. (Interview 9, item 34) |
|  | Prioritization could improve quality of studies | P4 | The quality of the protocols must improve. The kind of scientific thinking is simply often absurd. [...] So many technical mistakes are made (Interview 16, item 133). |
|  | Prioritization could avoid that trials are conducted with insufficient resources and expertise | P5 | [...] clinical studies with good valid data; that is also its own part of science, which requires the corresponding resources and expertise. (Interview 9, item 40)<br>They are then afraid that this will take something away from them, but I rather think the other way around: That I don't have to do at my clinic what five other clinics are doing in exactly the same way, which is a waste of time and money: Either we do it together or we don't do it at all, but I don't have to repeat the same research. (Interview 16, pos. 125) |
|  | Prioritization could reduce inefficiencies due to competitive recruitment | P6 | We are not competing with university hospital city against university hospital city, so where we have two university locations here, for example, or in university hospital city against Berlin, but in the end we are competing in a large international network. And of course it will not be the case that all 16 universities here have the same experts in the field of infectious diseases, especially since this is still a very small subject. But the few that are there have to pull together [...] the relevant stakeholders and can somehow moderate and manage them well, so that we at least speak with one voice to the public? (Interview 9, item 58) |
| Relevance | Prioritization could support the relevance of clinical studies | P7 | [...] this is indeed a problem and I would have wished that the national side would have coordinated this and said that these and the studies have priority, because they really bring us forward and not the Xth study, which examines something, but now really does not bring a solution in the sense of the pandemic. That would actually have been helpful. (Interview 2, item 31)<br>That has decreased over time, but when Covid really took off - in March, April, May of last year - we actually had several inquiries per day. And we actually looked at the projects and first considered, is it a scientifically meaningful concept? Is it also clinically reasonable for the patient? And is there a high probability that it will lead to an outcome that makes sense for the patient or for the research into the pandemic? [...] I would say in the order, scientific and clinical meaningfulness, secondly resources at the clinic and thirdly then also, what does it<br>perhaps bring in terms of third-party funding, in that order. (Interview 8, item 28) |
|  | Prioritization could identify patient-oriented knowledge gaps | P8 | So for the Covid patients who were in the ICU, ventilation studies were key. How do I ventilate? Zero funding, 0.0 funding from anybody. That means you'll find 0 ventilator studies. There aren't any. But for the person who is in the ICU, it's the most important thing. When do I start ECMO? How long do I blow oxygen? How much hypoxia can I tolerate? In the end-stage area, there are 0 studies, there are only case reports. (Interview 4, item 61) |
|  | Prioritization should include the allocation of biomaterials | P9 | Of course, that's always a good question: Is it necessary and right? And you have to be careful how much material is given to whom? Then the patient is already left out in the cold, so to speak. The material is there, the data are there, and then of course you have to look, with this limited material, "What research do I see as so important that it may be used for this?" (Interview 16, pos. 35) |
| Lack of explicit prioritization |  | P10 | It's [inquiries to participate in or recruit for clinical trials] completely unstructured. You are offered something, you don't have anything, and then you just take that, and sometimes there are actually overlaps. So that's problematic: We have several studies that are aimed at the same patient population and that's a problem (interview 5)<br>We have often said that it makes no sense to do this study alone: Either we leave it or we attach ourselves to a center elsewhere. (Interview 8, item 30) |
| Potential response strategies for prioritization |  |  |  |
| Central coordination and collaboration | National board | P11 | Well, of course, the one from which one hopes the most by reading in how really just the data situation was best. But of course one must say, here it would have been actually helpful, times in the past said, if a board had formed very fast, which would have had quasi really time to familiarize itself deeply. Maybe even really nationwide and would have said: "These are really the best candidates now. These are the ones we're counting on." So ultimately through our own research. And then: How feasible is that? Well, there are still a lot of inquiries. (Interview 11, item 22) |
|  | National cooperation | P12 | And I actually believe that at the beginning of the pandemic in Germany, we were probably in a particularly poor position in international comparison due to our<br>very decentralized structures and also, I would say, due to the pride we have in our decentralized structures. (Interview 15, item 6)<br>We are not competing with university hospital city against university hospital city, so where we have two university locations here, for example, or in university hospital city against Berlin, but in the end we are competing in a large international network. And of course it will not be the case that all 16 universities here have the same experts in the field of infectious diseases, especially since this is still a very small subject. But the few that are there have to pull together and that is perhaps one of the merits of *Mr. X*, who is responsible for the national guideline, that he brings together the relevant stakeholders and can somehow moderate and manage them well, so that we at least speak with one voice to the public?. (Interview 9, item 58) |
|  | Central registry | P13 | I think it would be good if the studies - of course it has to be voluntary again, otherwise it won't work - if the individual scientists could report to some central office which studies they are planning. They are then afraid that this will take something away from them, but I rather think the other way around: That I don't have to do at my clinic what five other clinics are doing in exactly the same way, which is a waste of time and money: Either we do it together or we don't do it at all, but I don't have to repeat the same research. (Interview 16, pos. 125) |
|  | Consultation by Ad-hoc expert groups | P14 | Perhaps temporary competence clusters would be needed, depending on time and topic, where... By competence clusters, I don't mean new institutes, but ad hoc committees composed of various study stakeholders who now come from different institutes and prepare studies in a bundled way. (Interview 18, item 45) |
|  | Efficient study planning | P15 | But of course, if you wish for something and look to the future, then this process must also run much faster at the European level. We have also had international studies in some cases, and this is very, very laborious. This actually has to be coordinated. In the |
|  |  |  | U.S., there have been important approval studies in this regard, in which we in Europe could not even participate because we were too slow. There were studies in the UK in which we could not participate in mainland Europe either, because different conditions prevailed here. And what was also very unfortunate was that large WHO studies actually passed Germany by again, also because of this very latent and partly poorly coordinated process. (Interview 13, item 24) |
|  | Effective recruitment of special patient groups | P16 | But I think if you really want to generate 'unique' research results from Germany ... We have enough patients. We have 2.5 million patients, right? If you wanted to recruit 'unique' patients, you would have had to do it top-down. (Interview 4, item 57) |
|  | Management of conflict of interest | P17 | Sure, such a board would help, but how can such a board be neutral? (Interview 11, item 11)<br>Because of course, as soon as someone is involved in the studies, he is "biased". And if he is not involved, maybe he is not biased. That's not so easy, I think. But of course one could imagine something like a board leadership. So you could say that the management levels of such a committee plus two additional members or so, a few real scientists who have distinguished themselves in the field [...]. So they are, so to speak, deeply involved in the matter, but in the end they don't have any shares in the game. (Interview 11, item 33) |
| Study design | Multicentre study design | P18 | Maybe also, if we were more multicentric, you would have much faster results. (Interview 14, item 29)<br>There are three studies running in parallel[...]. And of course it would be helpful to bundle them, because the number of cases and the spectrum of those included are getting larger. The background of those included also depends on whether I am in a practice at the university hospital or in a peripheral hospital, but we were<br>not able to bring the participants together. (Interview 16, item 54)<br>We have often said that it makes no sense to do this study alone: Either we leave it or we attach ourselves to a center elsewhere. (Interview 8, item 30) |
|  | Adaptive study designs | P19 | So if they do, then they just need a fixed established therapy group, such as ACTT [Adaptive COVID-19 Treatment Trial], where a joint design is then developed under fixed coordination, but there must then also be a team behind it, because it can then take over protocol [...] and other tasks in the pandemic - we see all this even now with pharmaceutical sponsors. (Interview 9, item 38)<br>So I do miss studies with adaptive design that can be adapted to the appropriate products and situations. That is sometimes a problem, because it means that you also have to look at how far adaptive study designs are actually applied. (Interview 18, item 45) |
| Alignment with funding | Funding for priority studies | P20 | There is no funding because there is no pharma. No funding because there is no virology, no basis in it, but it is a purely clinical question. I would like to see a central board. That's where the questions would be brought in. Then they say, for us these are the 'main' questions that need to be answered, delegate it to the respective 'disease' PI - if I call him that - who has to set up a study protocol and then funding has to be poured in before everyone does it in their own little box. (Interview 4, item 63) |
|  | Calls for priority topics | P21 | Well, of course, the one from which one hopes the most by reading in how really just the data situation was best. But of course one must say, here it would have been actually helpful, times in the past said, if a board had formed very fast, which would have had quasi really time to familiarize itself deeply. Maybe even really nationwide and would have said: "These are really the best candidates now. These are the ones we're counting on." So ultimately through our own research. And then: How feasible is that? Well, there are still a lot of inquiries. (Interview 11, item 22) |
|  | Efficient funding procedures | P22 | Far from ethics, it is obvious that the funding system in Germany is in no way prepared for a pandemic. In March, 150 million euros were thrown into the middle and people said, "Well, now we have to understand this<br>disease very quickly." And I think in December, the first euro flowed. And it wasn't because people didn't feel like drawing down the money, but because the mills have to grind in a very, very complicated way, due to legal requirements. (Interview 1, item 21)<br>The financial support is also so incredibly complicated and it takes an incredible amount of time. You are also held up so much by all these bureaucratic things that are behind it. And that is something that is a really big problem for a pandemic, where speed really counts. We received a lot of funding that year, also for clinical studies, but the organization of these is incredibly time-consuming. (Interview 10, item 20) |
|  | Fair funding procedures | P23 | I would have liked it to be similar to the money. If you want public funding now, you can't do it without *clinic X*. It got the big money. They got the big money and then you have to make sure that you get a little bit of *clinic X*, right? And [I] find it disastrous that the money went to virologists and the virologist primarily distributed it to other virologists. So the clinicians, they are completely ... if we didn't have access via pharma to study credit, we wouldn't have done any research at Covid as clinicians, but would have degenerated as bio-material suppliers for virologists. And then, of course, you're super interested if you know you're going to deliver and slave away all day and never get considered anyway. And I thought it was a pity that all the 'public grants' are controlled by *clinic X*, right? [...] No, that is due to the pandemic. *Mr. X* gets the whole thing and he distributes it. And for innovative clinical ideas you don't get any money from public funding, you have to approach the pharmaceutical industry, right? (Interview 4, item 18-20) |
|  | Sustainable funding | P24 | I think that a lot of the disruption came from the fact that, first of all, everyone wanted to contribute something to this global issue, but also that we nowadays finance a large part of our scientific work only through short-term third-party funding, right? So now, I have 31 female collaborators with my research group and not one of them is tenured, including myself. So, we all 32 work on temporary contracts and all on competitively acquired third-party funds. (Interview 15, item 6) |
| Pandemic policy on use/ownership of patient data | Agreement for secondary data use | P25 | Okay, so if we also want to participate scientifically here and above all we simply realized that something is now coming our way and we actually want to learn immediately while 'doing'. So the goal is that we don't say like we usually do, we're going<br>to collect some data now and evaluate that in a year and have our study protocol, our grant program or something, but actually more or less every week we actually want to prefer to evaluate what we learned from the last week because it's just so new, right, and coming so fast. (Interview 15, item 4)<br>Intervention study versus just data summary. And that's where I just come first to the question of having and using data from patients for later and also bio-materials. And I have to say, for this part my impression at Covid-19 was that the interest for the general public is so high that it would be helpful to define what is generally possible for the general public at a high level and does not require consent. (Interview 11, item 4) |
|  | Regulation on data use and access | P26 | Who has data sovereignty? Do I give away my data? Am I still considered enough there? So in an instant, everybody gets this feeling, "I'm giving something away. Am I possibly giving away my rights there?" And that somehow seems to be more possible in this NHS [*National Health Service*; Authority in UK], that the individual NHS doctor in the system - even if it's not China now - somehow functions better and immediately has the feeling that something is being taken away from him. (Interview 11, item 20) |
| Professionalisation of clinical research | Competence in clinical research | P27 | This is primarily a political question: How do we deal with the clinical trial landscape in Germany, and my main point of criticism is that there is not only a lack of a corresponding group - that is only the symptom of it - but it is simply assumed, especially by the very fundamentally oriented key opinion leaders, that anyone can conduct clinical trials, and that is simply fundamentally wrong in my eyes. (Interview 9, item 40)<br>That's perhaps part of the problem, that now people are doing clinical research who didn't really do clinical research in the past and then the collaborations don't go smoothly. (Interview 16, pos. 30)<br>And I realize that not every clinic can have an advanced training program for advanced studies, but there would have to be an institutional framework where younger physician colleagues experience a curricular element to then effectively set things up themselves afterwards. (Interview 18, item 24)<br>So if I could wish for something, and I want to turn it around, then it would be that the<br>university policy but also the social policy recognizes that clinical studies for high-quality requirements need their own scientific basis, the centers must be provided accordingly, especially in the field of infectious diseases or if you look at it more abstractly, in all German centers for health research. (Interview 9, pos. 58)<br>It's interesting to see who is conducting clinical studies now, and who is partially discontinuing them, so we once initiated studies here that never went into recruitment, which is of course simply a cost factor for everyone involved. (Interview 9, item 38) |
|  | Fair play among scientists | P28 | So if there is a collaborative body and if you try as an institution to make the research as efficient as possible in terms of the outcome, but at the same time in terms of the study participants to keep it as ethical as possible and also little resilient, then in the end you also have to look that there is also a fair play among the scientists. (Interview 8, item 4) |

## References

1. Janiaud P, Axfors C, Van’t Hooft J, Saccilotto R, Agarwal A, Appenzeller-Herzog C, Contopoulos-Ioannidis DG, Danchev V, Dirnagl U, Ewald H et al: The worldwide clinical trial research response to the COVID-19 pandemic - the first 100 days. F1000Res 2020, 9:1193.

2. Faust A, Sierawska A, Kruger K, Wisgalla A, Hasford J, Strech D: Challenges and proposed solutions in making clinical research on COVID-19 ethical: a status quo analysis across German research ethics committees. BMC Med Ethics 2021, 22(1):96.

3. Bigatello LM, George E, Hurford WE: Ethical considerations for research in critically ill patients. Crit Care Med 2003, 31(3 Suppl):S178-181.

4. Silverman H: Protecting vulnerable research subjects in critical care trials: enhancing the informed consent process and recommendations for safeguards. Ann Intensive Care 2011, 1(1):8.

5. Hirt J, Rasadurai A, Briel M, Düblin P, Janiaud P, Hemkens L: Clinical trial research on COVID-19 in Germany ? a systematic analysis [version 1; peer review: 1 approved]. F1000Research 2021, 10(913).

6. Pearson H: How Covid Broke the Evidence Pipeline. Nature 2021, 593(7858):182–185.

7. van der Graaf R, Hoogerwerf M-A, de Vries MC: The ethics of deferred consent in times of pandemics. Nature Medicine 2020, 26(9):1328–1330.

8. Meyer MN, Gelinas L, Bierer BE, Hull SC, Joffe S, Magnus D, Mohapatra S, Sharp RR, Spector-Bagdady K, Sugarman J et al: An ethics framework for consolidating and prioritizing COVID-19 clinical trials. Clinical Trials 2021, 18(2):226–233.

9. Mayring P: Qualitative Inhaltsanalyse : Grundlagen und Techniken, 12., überarbeitete Auflage edn. Weinheim; Basel: Beltz; 2015.

10. RECOVERY Trial [https://www.recoverytrial.net/]

11. BIO COVID-19 Therapeutic Development Tracker; https://www.bio.org/policy/human-health/vaccines-biodefense/coronavirus/pipeline-tracker

12. Singh S, Cadigan RJ, Moodley K: Challenges to biobanking in LMICs during COVID-19: time to reconceptualise research ethics guidance for pandemics and public health emergencies? J Med Ethics 2022, 48(7):466–471.

13. The RECOVERY Collaborative Group: Dexamethasone in Hospitalized Patients with Covid-19. New England Journal of Medicine 2020, 384(8):693–704.

14. Netzwerk Universitätsmedizin [https://www.netzwerk-universitaetsmedizin.de/]

15. Meerpohl JJ, Voigt-Radloff, S., Rueschemeyer, G., Balzer, F., Benstoem, C., Binder, H., Boeker, M., Burns, J., Dirnagl, U., Featherstone, R., Falk, F., Grundmann, H., Hengel, H., Kempf, V., Kern, W., Kranke, P., Laudi, S., Lieb, K., Maun, A., Mavergames, C., Metzendorf, M.-I., Nothacker, M., Schmaderer, C., Schmucker, C., Schwingshackl, L., Skoetz, N., Steckelberg, A., Stegemann, M., Strech, D., Moerer, O., Rehfuess, E., Spies, C.: CEOsys: creating an ecosystem for COVID-19 evidence. In: Collaborating in response to COVID-19: editorial and methods initiatives across Cochrane. Edited by Rev. CDS, vol. 12; 2020:9–11.

16. Nationales Pandemie Kohorten Netz [https://napkon.de/]

17. CODEX: COVID-19 Data Exchange Platform [https://www.medizininformatik-initiative.de/de/use-cases/codex-covid-19-data-exchange-platform]

18. Prokosch HU, Bahls T, Bialke M, Eils J, Fegeler C, Gruendner J, Haarbrandt B, Hampf C, Hoffmann W, Hund H et al: The COVID-19 Data Exchange Platform of the German University Medicine. Stud Health Technol Inform 2022, 294:674–678.

19. Ständiger Arbeitskreis der Kompetenz- und Behandlungszentren für Krankheiten durch hochpathogene Erreger [https://www.rki.de/DE/Content/Kommissionen/Stakob/Stakob_node.html]

20. Jha A, Lin L, Short SM, Argentini G, Gamhewage G, Savoia E: Integrating emergency risk communication (ERC) into the public health system response: Systematic review of literature to aid formulation of the 2017 WHO Guideline for ERC policy and practice. PLoS One 2018, 13(10):e0205555.

